# Maternal exposure to stress and child risk of obesity: a cross-sectional study of women and their children aged 5-15 years living in a deprived urban Peruvian community

**DOI:** 10.64898/2026.07.06.26355339

**Authors:** Emeline Rougeaux, Mary Fewtrell, Antonio Bernabé-Ortiz, Chen Song, Simon Eaton, Jonathan CK Wells, Edward Fottrell

**Author notes:** Corresponding author: Emeline Rougeaux.

## Abstract

**Objectives:** Maternal stress during childhood may contribute to childhood obesity through effects on parenting, parent–child relationships, or children’s stress responses. However, evidence is largely based on maternal self-reported stress, while studies using physiological measures like cortisol have found inconsistent associations. Most studies have been conducted in high-income settings, limiting generalisability. Using psychological and physiological stress markers, this cross-sectional study evaluates maternal stress exposures and child risk of obesity in 475 pairs of Peruvian women and their children, age 5-15 years, in a disadvantaged urban area.

**Methods:** Maternal stress exposures included mental distress (12-item General Health Questionnaire categorising moderate/high vs no/low distress) and allostatic load (AL; lower/moderate/higher AL assessed from Latent Profile Analysis of hair cortisol, BMI, waist circumference and blood pressure). Child outcomes included BMI and waist circumference-for-age z scores (BAZ and WCAZ). Linear regression analyses were conducted, adjusting for confounders.

**Results:** Versus mothers with no/low distress, those with moderate/high distress had children with 0.40 (95% CI: −0.66,−0.13) and 0.32 lower (−0.53,−0.11) child BAZ and WCAZ respectively. Versus lower AL mothers, moderate AL mothers had children with 1.15 (0.41,1.88) and 0.74 (0.20,1.28) greater BAZ and WCAZ while higher AL mothers had children with 1.43 (0.95,1.92) and 0.91 (0.50,1.32) greater BAZ and WCAZ respectively. Higher maternal cortisol independently predicted child adiposity.

**Conclusions:** Children of mothers with higher AL were at greater risk of overweight or obesity, which may add to the rising burdens of non-communicable diseases in resource-constrained settings as well as related social, economic, and public health costs.

## 1. INTRODUCTION

Since the 1990s, the prevalence of obesity in low- and middle-income countries (LMICs) has steadily increased (Zhou et al., 2026). While obesity in many LMICs is still concentrated in those with higher levels of income, we have seen a decrease in socio-economic inequalities driven by a faster rise in obesity in lower income groups (Templin et al., 2019). The most recent Peruvian report (2017-18) suggests over a third of children over 5 years have overweight or obesity and, in metropolitan Lima specifically, 45% of 6–13-year-olds and 35% of 12-17-year-olds are estimated to be living with overweight or obesity (UNICEF, 2023). Preventing childhood obesity is particularly important as it often persists into adulthood and increases risk of other NCDs and health conditions across the lifecourse, with considerable social and financial costs, particularly in LMICs (N. D. Ford, S. A. Patel, & K. M. Narayan, 2017; García-Morales et al., 2024; Kazibwe, Tran, & Annerstedt, 2021).

Childhood obesity is a complex problem with numerous risk factors spanning genetics, parental obesity, socio-economic circumstances, nutrition, lifestyle, environmental contaminants, early life conditions, and maternal factors (N. D. Ford et al., 2017; Shaban Mohamed et al., 2022). In childhood, studies are increasingly showing that parental stress may increase risks of child obesity through its influence on parenting, child-parent relationships and the child’s own stress and/or emotional response, in turn influencing the child’s eating, sleeping, sedentary behaviour, physical activity and fat deposition (O’Connor et al., 2017; Parks et al., 2012; Pereira et al., 2025; Pervanidou & Chrousos, 2012; Tate, Wood, Liao, & Dunton, 2015).

Stress can be defined as “the perceived or anticipated inability to successfully cope with situations that are not predictable or controllable” (Dorsey, Scherer, Eckhoff, & Furberg, 2022). Markers of stress include subjective measures that describe the lived experiences of stress, and objective measures that assess biological responses (Dorsey et al., 2022). The human stress response is complex and broadly involves a brain response that triggers the nervous system, glucocorticoid hormone secretion by the adrenal system, changes in organ function, and behaviour to face or evade threats (Charmandari, Tsigos, & Chrousos, 2005). When stress is prolonged or chronic, the response can become maladaptive, leading to elevated cortisol, inflammation and alterations in regulatory systems (Guidi, Lucente, Sonino, & Fava, 2020). Allostatic load describes the cumulative physiological burden of stress by bringing together measures of the primary mediators of the stress response (i.e. chemical messengers such as cortisol and epinephrine) and secondary outcomes (i.e. the cumulative effects on tissues and organs in response to the primary mediators such as inflammation markers, blood pressure, cholesterol, and body mass index) (Guidi et al., 2020; Knezevic, Nenic, Milanovic, & Knezevic, 2023; McEwen & Stellar, 1993).

Few studies from LMICs have explored associations between maternal stress and child obesity-related outcomes, despite these settings having a high burden of stress risk factors (e.g. poverty, pollution, noise, ill-health, natural disasters, and violent conflict) and increasing child obesity (Adam, 2023; Amsalem et al., 2025; Brisson et al., 2020; N. D. Ford et al., 2017; Gruebner et al., 2017; Lund et al., 2010). Furthermore, to our knowledge, none have done so using physiological measures like cortisol or composite measures of allostatic load. While studies of stress most often use psychological measures, these show weaker and less consistent effects than physiological measures – likely due to the greater influence of contextual factors (Gu, Lei, Yao, Chen, & Liu, 2025).

A systematic review and meta-analysis found that higher maternal psychological stress during childhood - including self-reported stress and anxiety, depression and mental distress - was generally associated with higher child BMI, particularly after infancy and when the child also experienced stress (Tate et al., 2015). However, the only study included in the review from a LMIC (Brazil) found the opposite (Rondó, Rezende, Lemos, & Pereira, 2013), while a subsequent study from Ethiopia found no association (Biadgilign, Mgutshini, Deribew, Gelaye, & Memiah, 2023). Furthermore, two studies from Greece and Germany which explored both psychological and physiological measures of parental/maternal stress, found positive associations only for anxiety and depression but not self-perceived stress and cortisol (Braig et al., 2023; Stymfaliadi et al., 2026).

Studies have shown that living in urban areas is associated with greater self-perceived stress and a more reactive physiological stress response (Gruebner et al., 2017; Lederbogen et al., 2011; Li, Ruan, Kang, & Rong, 2022; Lund et al., 2010; Steinheuser, Ackermann, Schönfeld, & Schwabe, 2014). Women may also experience greater stress than men due to differences in stress responsivity and social factors such as poorer access to financial resources, greater burdens of care, and higher exposure to intimate partner violence (Bhan, Rao, & Raj, 2020; Hazel & Kleyman, 2020; Kudielka & Kirschbaum, 2005).

Villa El Salvador is a district Peru’s capital, Lima, and is characterized by high levels of deprivation and a substantial migrant population (Castro & Riofrío, 1996; INEI, 2018a). This study aimed to assess associations between two indicators of maternal exposures to stress (mental distress and allostatic load) and child risk of obesity in Peruvian women and their children aged 5-15 years living in Villa El Salvador. We hypothesized that greater maternal stress, particularly mental distress, would be associated with greater child markers of obesity.

## 2. METHODS

### 2.1. Sample

The sample consisted of 490 pairs of women and their children aged 5-15 years living in the urban district of Villa El Salvador in the south of metropolitan Lima, Peru (**Figure 2; (Villa El Salvador Municipality)**) who took part in a study of internal migration and child health conducted in October 2023-March 2024.

**Figure 2.**
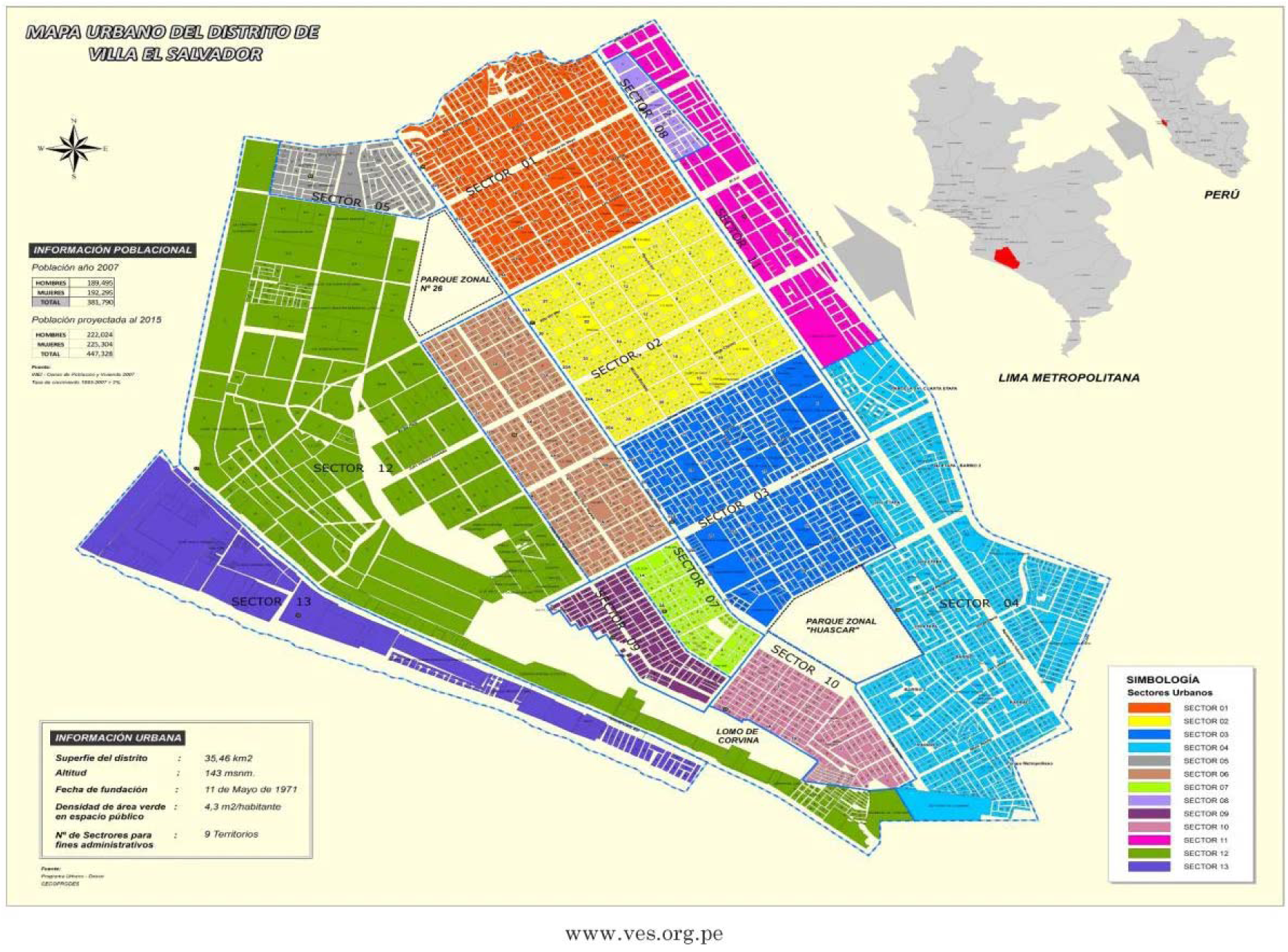
Map of Villa El Salvador

**Figure 2.**
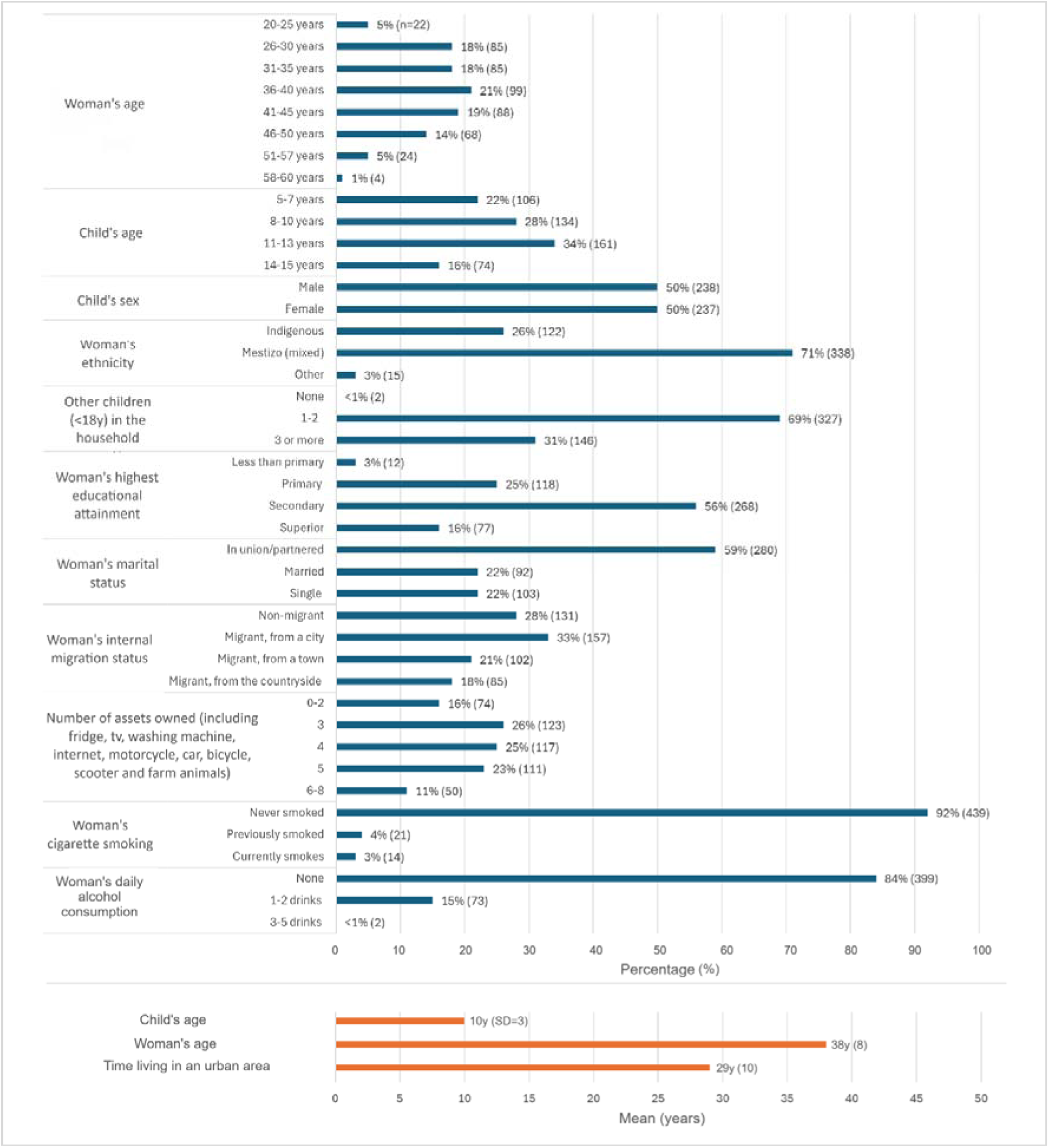
Characteristics of the analytic sample of women and children in Villa El Salvador (N=275)

Villa El Salvador was originally an informal settlement built in the 1970s in the desert outside Lima. The settlement then grew with the expansion of internal migration, particularly during the Shining Path conflict in the 1980s and 1990s (Thorp & Paredes, 2010). Subsequently, the Peruvian government formally recognized Villa El Salvador, allowing for political organization and access to amenities and services over time (Castro & Riofrío, 1996). It covers 35km^2^ and 2020 estimates suggest there are over 490,000 inhabitants (INEI, 2020). In 2020, it was the third poorest district in metropolitan Lima, with 20.2% of its population in monetary poverty; the study area was among the poorest neighbourhoods (INEI, 2018a).

The sample was collected use a non-random design, following both a convenience and quota sampling approach where households were selected based on ease of access and local knowledge of the area (due to limited resources and time and lack of existing census data) and a fixed target of non-migrant, urban-origin and rural-origin migrants. Details of sample size estimations can be found in **Appendix S1**. Where a mother had more than one child in the age range of interest, she was asked to select the oldest eligible child (contingent on the child providing informed assent, as described further on). Based on prior research and the knowledge and experience of the local team, we hypothesized older children/adolescents may be less interested in participating and/or less available, and hence should be prioritised when recruiting.

The fieldwork team went door-to-door in a residential area in the south of Villa El Salvador (sector 10 shown on **Figure 2**) inviting eligible mothers and their children to participate. Mothers provided separate informed consent for both their own, and their child’s participation and the child provided informed assent. Ethical approval was obtained from University College London (16813/001) and Universidad Peruana Cayetano Heredia (201838). The study consisted of an interview with the mother (using electronic tablets and a questionnaire on Open Data Kit) and physical measurements of the mother and child collected by four trained local fieldworkers in the mother and child’s home.

### 2.2. Measures

#### 2.2.1. Maternal exposure to stress

Evaluated using two measures reflecting the psychological and physiological impacts of stress: self-reported mental distress and measured allostatic load.

##### 2.2.1.1. Mental distress

The 12 question General Health Questionnaire (GHQ-12) designed by Goldberg was used (Goldberg et al., 1997). The GHQ-12 has been widely applied to assess mental wellbeing in migrant and non-migrant adults in Peru and validated in women of childbearing age in other Latin American settings (de Mola et al., 2012)(Rivas-Diez and Sánchez-López, 2014). The questions and response categories are shown in **Appendix S2**. These span different components of mental distress, including emotions (e.g. depression, anxiety), behaviours (e.g. self-confidence, concentration) and social functioning (e.g. enjoyment of activities).

For each question, there are four response options. These were scored using the bimodal system (where the first two received a score of 0 and the last two received a score of 1), resulting in a total score from 0 to 12 with higher values indicating higher distress. The GHQ-12 score was dichotomised as scores of 5 and over indicating moderate-high mental distress following other research in similar populations (Rivas-Diez and Sánchez-López, 2014, Loret de Mola et al., 2012).

##### 2.2.1.2. Maternal allostatic load

Five maternal biomarkers were available for measuring maternal allostatic load spanning neuroendocrine, metabolic and cardiovascular function. These were hair cortisol, BMI, waist circumference, systolic and diastolic blood pressure. Further detail of how these were combined is provided in the data analysis section.

###### Cortisol

Trained fieldworkers cut three locks of hair, each with a width approximately 2mm across, from three different locations on the posterior vertex of the mother’s head and then stored these together at room temperature in aluminium foil and sealed plastic bags. Following the protocol by Reid et al., a sample of 25mg of hair (3cm in length, reflecting approximately 3 months of cortisol) was selected from the scalp-end of the combined locks which was then washed twice in isopropanol, pulverized using five ceramic beads and a bead mill (Qiagen TissueLyser LT), and processed with methanol to extract its cortisol (Reid, Parker, Clemens, & Bristow, 2021). For this final extraction step 2mL of methanol was added to the powdered hair, then the mixture was shaken and incubated 24h at room temperature before being centrifuged and 1.4mL of methanol being extracted. This was then dried down using a vacuum centrifuge and tubes were stored at −80 Celsius until analysis.

On the day of analysis, samples were removed from the freezer and reconstituted using 0.15mL of cortisol immunoassay diluent buffer (Salimetrics) as per Reid et al.’s protocol (Reid et al., 2021). Cortisol levels were then measured in duplicate using Salimetrics Enzyme Linked Immunosorbent Assay kits following the manufacturer’s protocol (Salimetrics, 2021) and a plate reader (Tecan F200) at 450 nm.

Concentrations of cortisol were calculated, as recommended in the Salimetrics protocol, using a four-parameter logistic curve where background values were subtracted from all sample assay wells. After checking for erroneous or implausible values and cleaning the data, the mean concentration of duplicate samples was then converted from ug/dl to pg/mg using the following formula (Fenech et al., 2024):

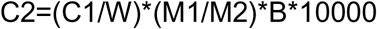

where C2 is the final mean concentration of hair cortisol in pg/mg, C1 is the mean concentration of hair cortisol in ug/dl from the assays, W is the weight of each hair sample (25mg), M1 is the volume of methanol added to the powdered hair (2ml), M2 is the volume of methanol extracted from M1 (1.4ml), B is the volume of assay buffer used to reconstitute the dry extract (0.15ml).

###### Body mass index

BMI was calculated as weight/height^2^ using means of two weight and height measurements taken during the home visit using Camry EB9013 electric scales (Camry Electronic Ltd, Hong Kong, China) and an Avanutri 312 portable height board (Avanutri, Tres Rios, Brazil) respectively.

For the descriptive analyses BMI categories were also created as healthy BMI (18.5 to <25 kg/m^2^), Overweight (BMI ≥25 to <30 kg/m^2^), and Obesity (BMI ≥30 kg/m^2^) (Weir and Jan, 2023).

###### Waist circumference

Waist circumference was measured twice at the midpoint between the bottom of the lowest rib and the top of the hip bone to the nearest millimetre using a Seca 201 measuring tape (Seca, Hamburg, Germany) and estimated from the mean of the two measurements.

For the descriptive analyses, central obesity was also defined a waist circumference >80cm (versus ≤80cm or no central obesity) (Alberti et al., 2009).

###### Blood pressure

Blood pressure was measured three times and at least 1 minute apart on the left arm using an Omron HEM 780 digital monitor and an appropriately sized cuff. The mother was in seated position with her forearm extended on a table in front of her, with the cuff placed on the upper arm approximately at heart level, and rested for five minutes prior to starting. The final measurements of diastolic and systolic blood pressure were estimated as the mean of the last two of the three measurements for each.

For the descriptive analyses, elevated blood pressure was also defined as elevated diastolic blood pressure (≥80 mmHg) and/or elevated systolic blood pressure (≥120 mmHg) (Whelton Paul et al., 2018).

##### 2.2.2.2. Child adiposity markers

The outcomes for children were waist circumference and BMI adjusted for age and sex.

###### Waist circumference

Waist circumference was measured in centimetres using a Seca 201 measuring tape (Seca, Hamburg, Germany) and using the same protocol as for the mother. Mean waist circumference was then estimated from the two measurements taken. This was then used to estimate internal z scores standardized for the child’s sex and age using the LMS method via the LMSGrowth application (Cole & Green, 1992). The result is referred to as waist circumference-for-age internal z score (WCAZ).

For descriptive analyses, WCAZ was dichotomised as being >90^th^ percentile (or >1.28 standard deviations [SD]), indicating a greater risk of central obesity (Xi et al., 2020).

###### Body Mass Index

BMI was calculated using height and weight as height/weight^2^. Height was measured standing and in centimetres using an Avanutri 312 portable height board (Avanutri, Tres Rios, Brazil). The two measurements of height taken from the child were then averaged. Weight also was measured twice and in kilograms using Camry EB9013 electronic scales (Camry Electronic Ltd, Hong Kong, China) and then averaged. BMI was transformed into z scores using WHO 2007 reference data for children aged 5-19 years with the Stata statistical software zanthro package, obtaining BMI-for-age z scores or BAZ (S. I. Vidmar, Cole, & Pan, 2013).

For descriptive analyses, categories of child BMI (thinness, healthy weight, overweight and obesity) were also estimated using the Stata zanthro package mentioned previously and age and sex standardized international cut-offs for children aged 2 to 18 years from Cole et al. and the International Obesity Taskforce (S. Vidmar, Carlin, Hesketh, & Cole, 2004).

#### 2.2.3. Covariates

Variables were initially selected as potential confounders in the association of maternal stress and child adiposity if they were associated with women’s stress, allostatic load, and/or mental distress and child adiposity-related outcomes in previous literature on similar populations and deemed not to be on the causal pathway (Chamik, Viswanathan, Gedeon, & Bovet, 2018; Christensen et al., 2018; N. D. Ford, S. A. Patel, & K. M. V. Narayan, 2017; Harkness & Hayden, 2020; Hernández-Vásquez, Rojas-Roque, Vargas-Fernández, & Bendezu-Quispe, 2020):

- Mother’s age (in years, categorized as 20-25, 26-30, 31-35, 36-40, 41-45, 46-50, 51-57, 56-60 years for the descriptive analyses)
- Mother’s migration status and type of previous residence (self-reported and categorized as non-migrant, city migrant, town migrant, countryside migrant)
- Mother’s ethnicity (self-reported and categorized as indigenous, mestizo or other)
- Maternal number of years lived in an urban area
- Child’s age (in years, categorized as 5-7, 8-10, 11-13, and 14-15 years for the descriptive analyses)
- Child sex (male/female)
- Maternal smoking (never smoked, previously smoked, or currently smokes cigarettes)
- Maternal daily alcohol consumption (none, 1-2 drinks, 3-5 drinks)
- Household assets (including fridge, tv, washing machine, internet, motorcycle, car, bicycle, scooter, and farm animals and categorized as 0-2, 3, 4, 5, 6, or 7-8 assets)
- Maternal education (less than primary, primary, secondary or superior)
- Other children (<18 years) living in the households (none, 1-2, 3+).

Additionally, for descriptive purposes, child stunting prevalence was estimated as height-for-age z score (HAZ) below 2 SDs of the reference median (FANTA, 2018). HAZ was estimated from average height (as defined previously) transformed into z scores using WHO 2007 reference data for children aged 5-19 years with the Stata statistical software zanthro package (S. I. Vidmar et al., 2013).

### 2.3. Statistical analyses

To create the measure of maternal AL, we conducted latent profile analysis by using a generalized structural equation model with a latent classes option and the five maternal biomarkers described previously. Compared to more traditional approaches, such as identifying a high-risk quartile based on the sample distribution, newer approaches like latent class and latent profile analyses provide a superior approach as biomarkers can contribute differently to AL and interact with each other as they would biologically (Carbone, Clift, & Alexander, 2022). Additionally, the use of latent profile analysis allows the inclusion of continuous measures, unlike latent class analysis and traditional approaches that only allow dichotomized variables.

Correlations were first assessed between the AL biomarkers; Pearson correlation coefficients and p-values are provided in the Supporting Information **Table S1**. For the latent profile analysis, a series of models were examined ranging from 1 to 6 possible classes; a class invariant diagonal three-class model with robust standard errors was selected based on the fit statistics described and presented in **Appendix S3.** Class probabilities, representing the proportion of women in that class, and predicted means of each biomarker are provided in the results section for each of the three classes. The three classes reflected lower, moderate and higher AL with means of all biomarkers increasing across these from the lower to the higher AL class. Latent class prevalences are provided in the results. Predicted means for BMI, blood pressure, waist circumference and cortisol for the three-class model are provided in **Table S2** in the Supporting Information.

Descriptive statistics of the sample are provided in a table in the results as medians (and interquartile range), means (and S.E.) or percentages (*n*). Univariate associations between covariates identified as potential confounders and both maternal exposure to stress (allostatic load and mental distress) and maternal and child metabolic outcomes were assessed using Pearson correlation, paired t or ANOVA tests as appropriate. In univariate analyses of cortisol and child outcomes cortisol was further log-transformed to address the skewed distribution. In the latent profile analyses, cortisol was not transformed but skewness was considered in the model specifications.

Relationships between maternal stress predictor and child adiposity outcome variables were first assessed using scatter plots and other graphical representations. Where applicable, heteroskedasticity and linearity were then tested using standardized predicted versus standardized residual plots and Breusch-Pagan tests.

Using logistic regression, we assessed the quantitative association between the two maternal stress exposures (allostatic load and mental distress) and, using linear regression, we assessed the quantitative associations of each stress exposure in turn with the two child adiposity markers (as illustrated in Supporting Information **Figure S1**). The latter were adjusted for confounders and for the alternative stress exposure (i.e. mental distress in the regression model of allostatic load and child adiposity markers and allostatic load in the regression model of mental distress and child adiposity markers). Correlation matrices and Variance Inflation Factor regression post-estimation were used to test for multicollinearity among independent variables.

Analyses were also conducted to explore associations between individual items of the mental distress and AL measures and the two child health outcomes. These consisted of linear regression of:

- each item of the GHQ-12 on each child outcome (BMI and waist circumference) in turn, first as univariate associations, then adjusted for confounders,
- each of the maternal allostatic load biomarkers (maternal log cortisol, BMI, waist circumference, systolic and diastolic blood pressure) and the child health outcomes, first as univariate associations, then adjusted for each other and then adjusted for confounders.

Results are reported as regression coefficients and 95% confidence intervals (95% CIs) for linear regression analyses and odds ratios and 95% CIs for logistic regression analyses. All analyses were conducted in Stata 19.5 (StataCorp., 2021).

## 3. RESULTS

### 3.1. Sample characteristics

A total of 490 pairs of mothers and children provided data in the study. Item missingness was low, with one maternal record missing for urban length of residence, one for smoking, one for alcohol consumption, and one for ethnicity. Six mothers were missing mental distress score due to non-response in the GHQ-12. Three children were missing BMI and waist circumference, and three mothers were missing cortisol measurements due to errors in measurement and/or labelling. After excluding these missing data, an analytical sample of 475 pairs was obtained.

**Figure 2** shows characteristics of the study analytic sample. Mother’s age ranged from 20 to 60 years old, though most were between 26 and 50 years old. Approximately 30% of mothers had three or more children <18 years living in the household at the time of the study interview, including the child who participated in the study. Among the children included in the study, there was a higher percentage of children aged 10-15 years old than 5-9 years old but a similar percentage of males and females. Mothers were more likely to report identifying as being of mixed (Mestizo) ethnicity followed by indigenous (26%, primarily Quechua) and other (which included White, Black/afro-Peruvian, and other unspecified). More than half of mothers had completed secondary school (56%). Few mothers reported either not going to school at all (<1%) or completing preschool level only (2%) or completing a university degree (1%). Most mothers had a partner, being either married (22%) or not married but in a union (59%). Most mothers were migrants (72%). For migrants, a city was the most common previous place of residence, followed by a town and then the countryside. The average time spent in an urban area over the mother’s lifetime was 29 years. Sixteen percent of mothers reported owning only 0 to 2 assets and 26% 3 assets from a fridge, tv, washing machine, the internet, motorcycle, car, bicycle, scooter, and farm animals.

**Figure 3** summarises maternal stress exposure and maternal and child metabolic health outcomes. Mothers had a median level of hair cortisol of 9 pg/mg (interquartile range: 6 pg/mg) and a prevalence of moderate/high mental distress of 22%. Eighty four percent of mothers were living with overweight or obesity and 88% with central obesity. No mothers were classified as thin (BMI ≤18). Fourteen percent had elevated blood pressure. Thirty nine percent of women belonged to the lower AL class, 52% to the moderate AL class, and 9% to the higher AL class. Among their children 17% were at risk of general obesity and 11% of central obesity (waist circumference-for-age >90th percentile). Three percent of children had stunting.

**Figure 3.**
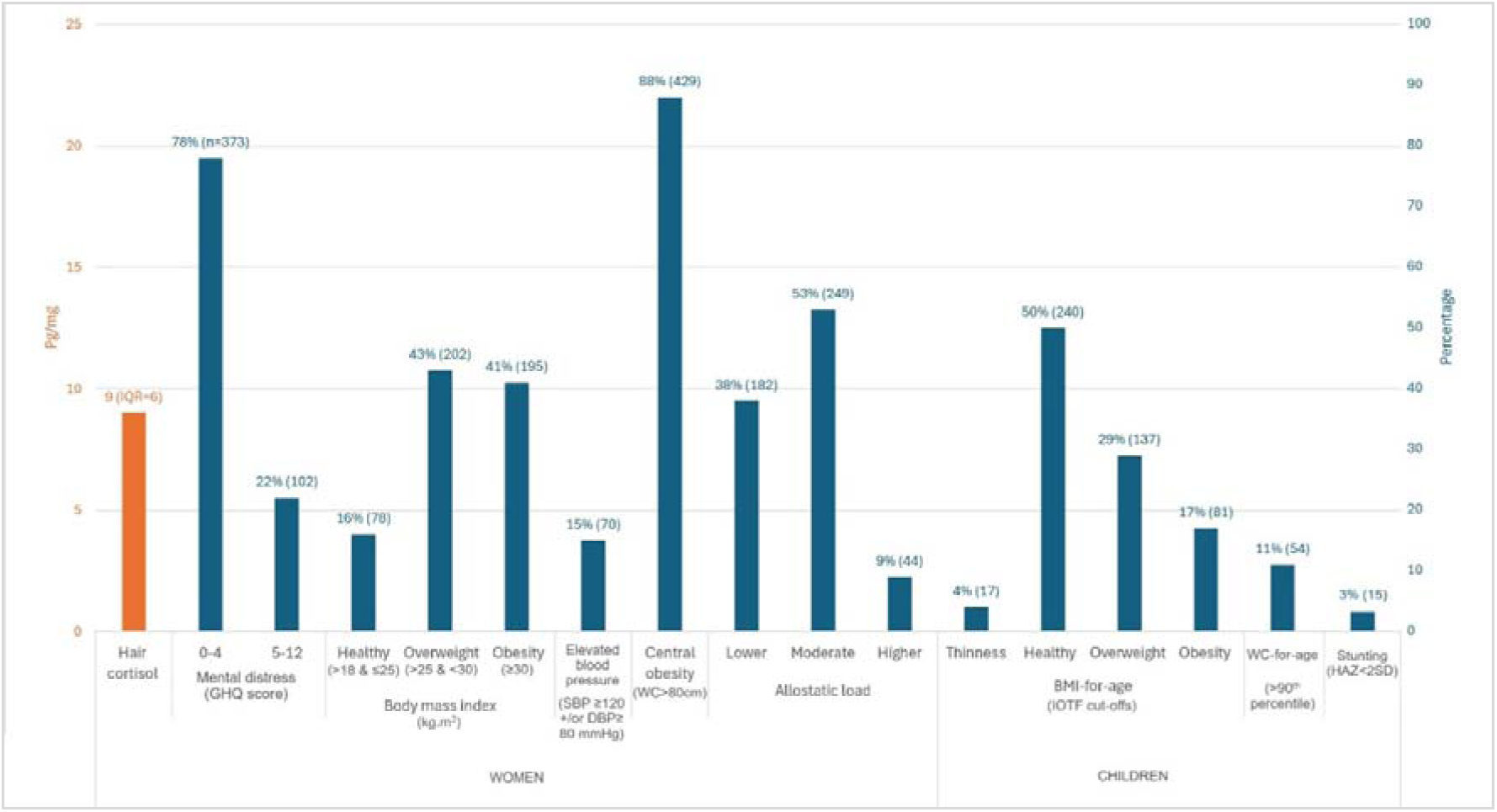
Health characteristics of women and children in the Villa El Salvador maternal migration child health study analytic sample (N=475) Footnotes: BMI=body mass index, GHQ=General Health Questionnaire, IQR=interquartile range, WC=waist circumference, SBP=systolic blood pressure, DBP=diastolic blood pressure, IOTF= International Obesity Task Force, HAZ=height-for-age z score, SD=standard deviation. BMI-for-age cut-offs and stunting are based on international cut-offs for children 2-18 years (FANTA 2018; Vidmar et al. 2013). Allostatic load was estimated from class predictions following Latent Profile Analysis of measures of cortisol, BMI, blood pressure and waist circumference.

### 3.2. Association of maternal exposures to stress and child health outcomes

Initial analyses indicated moderate to strong correlations between maternal BMI, waist circumference and blood pressure and weak correlations between hair cortisol and other markers (apart from diastolic blood pressure where there was little evidence of correlation) (Supporting Information, **Table S1**). Child BMI-for-age and waist circumference-for-age were also strongly correlated with each other (Supporting Information, **Table S3**).

Associations between the two measures of maternal exposure to stress were then assessed. Compared to women in the lower AL group, those with moderate AL had 2.02 (1.21, 3.38) times the odds of having moderate/high mental distress but there was less evidence of an association for those with higher AL (OR: 1.42 [0.60, 3.37]) (Supporting Information, **Table S4**).

**Table 3** presents the results of the linear regression of maternal mental distress and the two child adiposity markers (BAZ and WCAZ). The results indicate a small negative association for both outcomes. After adjusting for confounders (child sex, age, maternal age, migration status, ethnicity, and years lived in an urban area), mothers with moderate/high mental distress had children with 0.31 (95% CI: −0.59, −0.03) lower z scores of BMI-for-age and 0.25 (−0.47, −0.04) lower z scores for waist circumference-for-age. These differences were increased (to −0.40 [−0.66, −0.13] of BAZ and −0.32 [−0.53, −0.11] of WCAZ) when maternal AL was also included in the model (**Table 3**).

**Table 3.** Associations of maternal mental distress and child adiposity markers (N=275)

|  |  | Unadjusted<br>model<br>Coefficient<br>(95% CI) | Adjusted for<br>confounders†<br>Coefficient<br>(95% CI) | Adjusted for<br>confounders &<br>maternal<br>allostatic load<br>Coefficient<br>(95% CI) |
| --- | --- | --- | --- | --- |
| Child BMI-for-age<br>z-score (WHO) | No/low maternal mental<br>distress | Ref. | Ref. | Ref. |
|  | Moderate/high maternal<br>mental distress | -0.35<br>(-0.63, -0.07)* | -0.31<br>(-0.59, -0.03)* | -0.40<br>(-0.66, -0.13)* |
| Child waist<br>circumference-for-age z score<br>(internal) | No/low maternal mental<br>distress | Ref. | Ref. | Ref. |
|  | Moderate/high maternal<br>mental distress | -0.26<br>(-0.48, -0.05)* | -0.25<br>(-0.47, -0.04)* | -0.32<br>(-0.53, -0.11)* |

† Confounders include child sex, age, maternal age, migration status, ethnicity, and years lived in an urban area; \* p<0.05; BMI=body mass index, WHO=World Health Organization, CI=confidence interval. Maternal mental distress score was obtained from the 12 item General Health Questionnaire and dichotomised as scores of 5 and over indicating moderate/high distress. Allostatic load was estimated from class predictions following Latent Profile Analysis of measures of cortisol, BMI, blood pressure and waist circumference and categorized as lower, moderate and higher allostatic load.

For univariate analyses of the GHQ-12, all coefficients suggested negative associations with child BMI and waist circumference though effect sizes varied across these (Supporting Information, **Table S5**).

**Table 4** presents the results of the linear regression of maternal AL and the two child adiposity markers. Greater AL was positively associated with both BAZ and WCAZ, with larger effects sizes for BAZ. After adjusting for confounders and compared to mothers with lower AL, mothers with moderate and higher AL had children with a greater BMI-for-age of 0.63 (0.39, 0.87) and 1.28 (0.87, 1.69) z scores respectively and a greater waist circumference-for-age of 0.45 (0.26, 0.64) and 0.85 (0.53, 1.18) z scores respectively. When maternal mental distress was also adjusted for, effect sizes increased slightly across both outcomes (**Table 4**).

**Table 4.** Associations of maternal allostatic load and child adiposity markers (N=275)

|  |  | Unadjusted<br>model<br>Coefficient (95%<br>CI) | Adjusted for<br>confounders†<br>Coefficient (95%<br>CI) | Adjusted for<br>confounders &<br>maternal mental<br>distress<br>Coefficient (95% CI) |
| --- | --- | --- | --- | --- |
| Child BMI-for-age z-score<br>(WHO) | Lower AL | Ref. | Ref. | Ref. |
|  | Moderate AL | 0.58 (0.34, 0.81)* | 0.63 (0.39, 0.87)* | 0.67 (0.43, 0.91)* |
|  | Higher AL | 1.25 (0.84, 1.66)* | 1.28 (0.87, 1.69)* | 1.30 (0.87, 1.70)* |
| Child waist circumference-for-age z score (internal) | Lower AL | Ref. | Ref. | Ref. |
|  | Moderate AL | 0.43 (0.25, 0.62)* | 0.45 (0.26, 0.64)* | 0.49 (0.30, 0.67)* |
|  | Higher AL | 0.88 (0.57, 1.20)* | 0.85 (0.53, 1.18)* | 0.87 (0.55, 1.19)* |

† Confounders include child sex, age, maternal age, migration status, ethnicity, and years lived in an urban area;
\* p<0.05; AL=allostatic load, BMI=body mass index, WHO=World Health Organization, CI=confidence interval. Maternal AL categories were estimated from latent profile analysis of maternal measured hair cortisol, BMI, blood pressure, and waist circumference. Maternal mental distress score was obtained from the 12 item General Health Questionnaire and dichotomised as scores of 5 and over indicating moderate-high mental distress.

Univariate analyses of individual biomarkers of AL suggested positive associations of all five maternal biomarkers and both child outcomes. In the fully adjusted models, which included both confounders and all biomarkers, associations were primarily seen for a. maternal cortisol and BMI and child BMI (after adjusting for confounders and maternal waist circumference and blood pressure) and b. maternal cortisol and waist circumference and child waist circumference (after adjusting for confounders and maternal BMI and blood pressure) (Supporting Information**, Table S6**).

In these models we find that, an increase of 1 unit of maternal cortisol (log pg/mg) was associated with an increase of 0.18 (−0.02, 0.39) z scores of child BMI-for-age and 0.14 (−0.02, 0.30) z s scores of child waist circumference-for-age. Additionally, an increase of 1 kg.m^2^ of maternal BMI was associated with an increase of 0.07 (0.03, 0.12) z scores of children BMI-for-age (but not waist circumference-for-age) and an increase of 1 cm of maternal waist circumference was associated with an increase of 0.03 z scores of child waist circumference-for-age (**Table S6**).

## 4. DISCUSSION

### 4.1. Summary of findings and implications

Independent associations were found for each maternal stress exposure and both child adiposity markers but in opposite directions and with stronger effect sizes for AL than for mental distress. After considering both confounding and AL, mothers with moderate/high mental distress had children with lower z scores (of 0.40 for BMI and 0.32 for waist circumference) compared to mothers with no/low mental distress. In contrast, compared to mothers with lower AL, those with moderate and higher AL had children with greater z scores (of 0.67 and 1.30 for BMI and 0.49 and 0.87 for waist circumference for moderate and higher AL respectively). Further analyses with individual biomarkers of AL suggest that both maternal cortisol and markers of adiposity are independently associated with child markers of adiposity.

These findings suggest children of mothers with moderate and - in particular - higher AL are at risk of higher BMI and waist circumference. This may be associated with a greater risk of overweight or obesity, which is likely to persist into adulthood and increase their risks of developing associated health conditions like cardiovascular disease, hypertension and diabetes (Baird et al., 2017; Darsini, Hamidah, Notobroto, & Cahyono, 2020; S. I. Vidmar et al., 2013; Xi et al., 2020). Obesity and other NCDs impose significant social and financial burdens due to their effects on social functioning, quality of life, workforce productivity, and healthcare costs. These challenges are particularly pronounced in LMICs, due to high poverty rates and prevalence of communicable and other health conditions, limited access to health services, and inadequate financial risk protection (N. D. Ford et al., 2017; García-Morales et al., 2024; Kazibwe et al., 2021).

### 4.2. Comparison with other research

As described previously, Tate et al.’s systematic review showed an overall positive association between maternal self-reported psychological stress (which included measures of stress and mental health problems like depression and anxiety) and child obesity-related outcomes, though none of the studies included were from LMICs (Tate et al., 2015). More recent studies have had mixed findings, with a longitudinal study in Germany showing that maternal self-perceived stress in the first year of her child’s life was associated with greater child BMI up to 5 years but only in girls (Leppert et al., 2018), a cross-sectional study from Brazil finding a null or weak negative association with obesity in children aged 6-12 years (Pereira et al., 2025), and another longitudinal study from the USA showing no association with child BMI at 11-15 years (Dunton, Chu, Naya, Belcher, & Mason, 2021). Tate et al. also assessed whether the association differed when measures of stress included mental health problems and reported no difference compared to not including mental health problems. In contrast, Marco et al. only found a positive association between parental poor mental health and child obesity in half of the studies identified in their systematic review, all of which were of either mental distress or depression and only one of which was from a LMIC (Brazil) (Marco, Valério, Zanatti, & Gonçalves, 2020). Two UK studies report positive associations between maternal mental distress and child overweight/obesity at 3 and 5-11 years, while one from Ethiopia reports no association at 5-18 years (Biadgilign et al., 2023; Hope, Micali, Deighton, & Law, 2019; Ramasubramanian, Lane, & Rahman, 2013). Different study designs, sample sizes, confounders, types and ways of assessing stress exposures may explain the heterogeneity across studies.

One study has explored maternal AL in pregnancy, finding a positive association with child fat mass from birth to six years (Gyllenhammer et al., 2025) but to our knowledge, no studies have explored maternal AL during childhood and associations with child markers of adiposity or obesity. Among studies of individual biomarkers of AL and child obesity, two studies – one from Germany exploring maternal cortisol up to 12 months and child BMI at 3 years and another from Greece exploring parental cortisol and child BMI at 2-12 years– both found no association (Braig et al., 2023; Stymfaliadi et al., 2026). However, the first study assessed maternal salivary cortisol - which has been found to be a less reliable measure than hair cortisol (Ma et al., 2022) - and the second study assessed hair cortisol only in a sub-sample of 13 of 103 parents (Stymfaliadi et al., 2026). No associations were found with child BMI for maternal/parental self-reported stress, though the German study did find positive associations for maternal anxiety in girls and the Greek study for parental BMI, anxiety and depression (Braig et al., 2023; Stymfaliadi et al., 2026). Other markers of maternal AL such as inflammation and obesity, both in pregnancy and postnatally, have been associated with greater child BMI, waist circumference and/or obesity across several studies (Denizli, Capitano, & Kua, 2022; Romy Gaillard & Jaddoe, 2016; R. Gaillard, Rifas-Shiman, Perng, Oken, & Gillman, 2016; Wu et al., 2018).

### 4.3. Potential mechanisms

Most study women and children had lived in Villa El Salvador for much or all their lives and likely had long-term exposure to stresses associated with life in a deprived, densely populated urban informal area. Many women are also internal migrants of indigenous ethnicity, both associated with higher risks of poverty, discrimination and intimate partner violence – all of which may increase stress and poor mental and physical health (Cevallos, 2023; Kohrt, Barrueco, & Perez, 2015; Miller, LaFave, Marineau, Stephens, & Thorpe, 2021; Moreno & Oropesa, 2012; Starn, Kirk, Degregori, & Kirk, 2009; Wang, Li, Stanton, Fang, & medicine, 2010; Zhang, Li, Fang, & Xiong, 2009). Migrant and indigenous women are also more likely to have experienced the Shining Path conflict of the 1980-90s, which mainly affected indigenous Peruvians and led many to migrate to coastal informal settlements like Villa El Salvador (Thorp & Paredes, 2010).

Little research has explored stress among Latin American populations and particularly indigenous groups. A study on Tsimane’ hunter-forager-horticulturalists in Bolivia found no association between hair cortisol and adiposity within adults and a negative association within children (Bethancourt, Ulrich, Almeida, & Rosinger, 2021). Other research from Brazil found serum cortisol to be positively associated with central adiposity in urban but not rural areas (Freitas et al., 2018). This may suggest that rural and urban populations cope differently with stress and that stress may increase obesity risk most in obesogenic environments. However, previous research has shown dampened salivary cortisol in the Tsimane and another similar population in Papua New Guinea when compared to western industrialized populations (Nyberg, 2012; Urlacher, Liebert, & Konečná, 2018), echoing other studies showing blunted cortisol in adults who were exposed to adverse experiences in/since childhood in the Philippines and USA or women with major depression in Mexico (Adler, Gertler, Fernald, & Burke, 2005; Desantis, Kuzawa, & Adam, 2015; Kuras et al., 2017). It has been suggested this may be an adaptive mechanism to prevent the health effects of hypercortisolism in environments with high burdens of stress; though stressors like nutritional deprivation, infectious diseases, and high levels of physical activity may also directly reduce cortisol (Urlacher et al., 2018). Blunted cortisol may explain why hair cortisol was lower in our study compared to many others (Igboanugo, O’Connor, Zitoun, Ramezan, & Mielke, 2024). Comparisons across studies should however be made with caution due to the many sources of variability (e.g. tissue type, environmental exposures, health behaviours, laboratory methods) and the lack of reference data (Igboanugo et al., 2024)

Over 80% of Peru’s population is now urban with those of indigenous descent disproportionally living in poorer areas on the outskirts of cities (Del Popolo, Oyarce, Ribotta, & Rodríguez Vignoli, 2007; INEI, 2018b). With the shift away from rural traditional lifestyles towards more western urban patterns, we have seen increasing burdens of obesity and metabolic disease among indigenous groups, such as the Awajun in Peru and the Toba in northern Argentina (Lagranja, Phojanakong, Navarro, & Valeggia, 2015; Qasim et al., 2018; Tallman, Valdes-Velasquez, & Sanchez-Samaniego, 2022). Other research from Peru showed a 12% increased incidence of obesity among rural-urban Andean migrants for every 10 years of urban residence, with no change seen in non-migrant rural and urban comparators (Antiporta, Smeeth, Gilman, & Miranda, 2016). At the same time, we continue to see higher rates of undernutrition in indigenous children; the effects on metabolic capacity may mean a greater predisposition to obesity-related NCDs when individuals are then exposed to urban obesogenic environments and lifestyles (Torres-Roman, Urrunaga-Pastor, Avilez, Helguero-Santin, & Malaga, 2018; J. C. K. Wells, 2018). Lower health care coverage and greater incidence of illness likely explain greater undernutrition, stemming from persistent and systemic language and cultural barriers to health care and exclusionary practices which can be traced back to colonial times (Gatica-Domínguez, Mesenburg, Barros, & Victora, 2020; Serván-Mori & Meneses-Navarro, 2025). It has also been suggested that there may certain genetic adaptive traits among indigenous groups designed to survive historical food-scarce environments/periods and/or protect against the effects of disease which, in modern societies characterized by food abundance and low energy expenditure, may predispose to obesity and related NCDs – though this theory remains highly debated (Johnson et al., 2022; Qasim et al., 2018).

Parenting behaviours may explain links between maternal stress and child obesity (Tate et al., 2015). A USA study found parenting stress associated with greater fast-food consumption in both parents and their children (Bautista et al., 2023). Little research has examined parental stress and child ultra-processed food consumption elsewhere, though this may be particularly relevant in Latin America where such foods are cheap, widely available, and contribute to rising obesity (Matos, Adams, & Sabaté, 2021). Parenting under stress may be more authoritarian or permissive, leading to poorer child eating habits, disordered satiety, and weight gain (Gouveia, Canavarro, & Moreira, 2019). Higher anxiety or depression symptoms have also been linked to nonresponsive feeding practices, characterised by persuasive feeding and rewards for behaviour and/or eating, and greater child emotional eating (Sampige, Kuno, & Frankel, 2023). Parents’ own behaviours further influence those of their children; for example, emotional eating, physical activity and sleep are corelated between parents and children and may influence overweight risk (Dakanalis et al., 2023; Sampige et al., 2023) (Maia, Braz, Fernandes, Sarmento, & Machado-Rodrigues, 2025). While O’Connor et al. found no link between maternal stress and child diet in their systematic review, they reported consistent positive associations with lower physical activity and higher sedentary behaviour, suggesting these as key mechanisms (O’Connor et al., 2017).

These associations may depend on socio-economic contexts. Poverty has been shown to increase parental stress and negatively affect parenting and parent-child relationships (Ho et al., 2022). Berge et al. found that parents in food insecure households experiencing stress and low mood were more likely to use restrictive feeding practices and provide pre-prepared meals to their children than those in food-secure households (Berge et al., 2020). Associations between child eating habits and obesity risk have also been found to be stronger in more disadvantaged neighbourhoods (Tarro, Lahdenperä, Vahtera, Pentti, & Lagström, 2022). In contexts of poverty, many stresses occur at the household level and are likely experienced directly by the child, particularly as they get older. In Tate et al.’s review, associations between maternal psychological stress and obesity were strongest when the child had also experienced stress and when maternal stress and child obesity were assessed later in childhood (Tate et al., 2015). Older children may be more aware of family and community problems and have had a longer exposure, which may be explain mixed findings across studies.

In poor urban contexts, families are often exposed to environmental stresses such as noise and pollution, which may explain both AL in the mother and increased adiposity in the child (Corburn & Sverdlik, 2019; Gani et al., 2025). Studies show both noise and air pollution trigger physiological stress responses and may increase AL (Sivakumaran et al., 2022; Thomson, 2019; Walker, Brammer, Cherniack, Laden, & Cavallari, 2016). Air pollution can also directly impact child growth, which, as described previously, may influence later risk of obesity. In Peru, internal migrants in urban areas had 10-fold higher exposure to particulate matter 2.5 than non-migrants (Carrasco-Escobar, Schwarz, Miranda, & Benmarhnia, 2020; Sinharoy, Clasen, & Martorell, 2020). Pollution may impact child linear growth by increasing oxidative stress and systemic inflammation prenatally and risks of respiratory infection postnatally (Sinharoy et al., 2020). Systematic reviews further suggest a positive association between ambient air pollutant exposure and obesity risk in children and adults (Luo et al., 2024). Chronic noise exposure may also increase the obesity risk(An, Wang, Ashrafi, Yang, & Guan, 2018; Liang et al., 2022).

It is unclear why, unlike AL, maternal mental distress was negatively associated with child adiposity markers in our study in contrast with other studies discussed previously(Biadgilign et al., 2023; Hope et al., 2019; Ramasubramanian et al., 2013). However, these studies used the 20-item Self Reporting Questionnaire, which includes questions relating to depression, anxiety and physical symptoms and the Kessler-6, which focuses mainly on nervousness and depression. In contrast, the GHQ-12 used in our study includes questions relating to depression, anxiety and social functioning (see **Appendix S2**). Other studies have used the Perceived Stress Scale (such as (Dunton et al., 2021; Leppert et al., 2018; Pereira et al., 2025)), which includes questions on anxiety but not depression. Thus, studies could be assessing different aspects and/or effects of stress, which may explain heterogenous findings. In our study, all items of the GHQ-12 were negatively associated with child BMI and waist circumference, though coefficients varied suggesting that some items (e.g. losing confidence in oneself or feeling worthless) may be more important than others (e.g. difficulty concentrating or losing sleep over worry). Furthermore, the GHQ-12 captures recent maternal mental distress. Chronic or long-term mental distress may show different associations with AL and child adiposity markers. Longitudinal research is needed to clarify causal links between maternal stress, mental health, AL and child obesity risk.

Further research is also needed to understand how maternal distress is associated with child nutritional health, including potential undernutrition or disordered eating, as poor maternal mental health has been associated with increased risk of eating disorders in children (Barakat et al., 2023).

### 4.4. Strengths and limitations

The strengths of this study include a large sample of close to 500 women and their children, a wide age range for children from 5 to 15 years and measured physical variables in both the mother and child. In particular, the use of hair cortisol in the mother has allowed the assessment of average basal cortisol over a longer period (with 3 cm of hair indexing approximately 3 months of cortisol) and is not subject to fluctuations like saliva or urine cortisol measurements are (Wosu, Valdimarsdóttir, Shields, Williams, & Williams, 2013). Anthropometric and blood pressure measurements were collected by trained staff using protocols that minimized errors in measurements, for example through repeated measurements by the same fieldworker and with help from the mother as needed. Mental distress was assessed with a widely used and validated tool, which had also previously been used in other research in nearby settings in similar populations (Loret de Mola et al., 2012).

Limitations include the cross-sectional design. Though research suggests that maternal stress may increase the risk of child obesity, our study design limits our ability to infer any causality. There are also limitations relating to our measurement of exposure to stress. Notably we had no biomarkers of immune function or inflammation and therefore our measure of AL does not reflect all the different types of physiological responses and impacts of stress. We may have obtained different classes of AL with the inclusion of additional biomarkers and different associations with child outcomes. The cross-sectional design also meant we could not assess to what extent increased cortisol and blood pressure may also have resulted from having overweight or obesity (Foss & Dyrstad, 2011). As noted previously, mean hair cortisol in our sample also appeared lower than many other studies; while this may reflect blunted cortisol, it is possible that cortisol was reduced by hair treatments, for example many of the women in the sample appeared to have dyed hair (based on colour residues seen in many of the cortisol samples after hair processing), and by long-term storage (as the laboratory analyses were conducted 1-1.5 years after hair collection) (Hoffman, Karban, Benitez, Goodteacher, & Laudenslager, 2014; Huthsteiner, Finke, Peters, Klucken, & Stalder, 2025). Though BMI-for-age is widely used to assess child risk of obesity in epidemiological research, it has limitations as it does not differentiate between lean and fat mass and has been found to be a poor predictor of fat mass in younger children(Jonathan CK Wells, 2014). However, it has been strongly associated with fat mass in late childhood and adolescence and demonstrated high validity for measurement of high body fat in school children in Mexico (Marcos et al., 2025; Vanderwall, Randall Clark, Eickhoff, & Carrel, 2017). In other work comparing body composition (from four-component model and dual X-ray absorptiometry) and BMI in children, researchers found that while both fat mass and fat-free mass increased across quintiles of BMI z score in the arm, leg and trunk, fat mass had the steepest rate of increase (J. C. Wells et al., 2006). To avoid wrongly categorizing children, we opted for continuous z scores. We also included a secondary outcome, waist circumference-for-age, which has been found to strongly correlate with visceral adipose tissue in children (Brambilla et al., 2006) and which was found to strongly correlate with BMI-for-age in our study. Finally, it is possible that the measure of maternal distress may have some social desirability bias (e.g. underreporting due to stigma around mental health). Future longitudinal research is needed to compare psychological and physiological assessments of stress and assess the influence of different stressors, such as air pollution, noise, violence, poverty and discrimination, which may help shed light on causality and mechanisms of association.

## 5. CONCLUSION

In this sample of women and their children aged 5-15 years in a deprived urban community in Peru, greater maternal AL, a physiological marker of cumulative exposure to stress, was associated with higher child BMI and waist circumference. This suggests maternal long-term stress exposures may predispose children to obesity and related NCD risk. In contrast, recent maternal mental distress, a marker of short-term psychological stress, was negatively associated with child BMI and waist circumferences. A key limitation of the study is the cross-sectional design, warranting further longitudinal research to establish causal links between maternal stress exposures, mental health, AL and child growth and nutritional health. Interventions that reduce or offset maternal stress may still help lower maternal AL and child adiposity markers, and reduce the growing NCD burden in LMICs.

## LIST OF ABBREVIATIONS

DHS: Demographic and Health Surveys
LMICs: low- and middle-income countries
BMI: body mass index
BAZ: BMI-for-age z scores
WCAZ: waist circumference-for-age z scores
HAZ: height-for-age z scores
WHO: World Health Organization
SD: standard deviations
AL: allostatic load
GHQ: General Health Questionnaire
IQR: interquartile range

## DECLARATIONS

### Ethics

Ethical approval was obtained from University College London (16813/001) and Universidad Peruana Cayetano Heredia (201838).

### Availability of data and materials

Data and study materials are available upon reasonable request from the authors.

## Authors contributions

ER designed the primary research study with support from ABO, JW, and MF; ER and ABO prepared study materials and ABO oversaw data collection. ER identified the research question, carried out the statistical analyses and drafted the paper. ER carried out the laboratory analyses with training from CS and under the supervision of SE. CS, SE, JW, MF, ABO, and EF contributed towards shaping the research question and the analyses and read and approved the final manuscript.

## Data Availability

Data and study materials are available upon reasonable request from the authors.

## Acknowledgements

We would like to express our gratitude to Dr.Elisa Ruiz Burga, Lilia Cabrera Rojo, Carmen Quinteros Reyes, and the fieldwork team for their diligence and assistance in carrying out the research project, as well as, above all, to the women and children of Villa El Salvador for their invaluable involvement.

## Disclosures

### Use of Artificial Intelligence

The authors made no use of any artificial intelligence (AI) tools in the writing, editing, or data analysis of this manuscript.

