## Supporting Information for "Maternal exposure to stress and child risk of obesity: a cross-sectional study of women and their children aged 5-15 years living in a deprived urban Peruvian community"

#### Appendix S1: Primary study sample size calculations

The primary study was originally designed to compare growth and nutritional health in children of urban non-migrant, migrant of rural origin and migrant of urban origin women. An ideal sample size of 600 mothers (including 150 urban-origin migrants and 300 rural-origin migrants) and their children aged 5-15 years was estimated based on effect sizes in other existing studies in Peru as described below.

Calculations of effect size were made using Demographic & Health Survey (DHS) data for Peru used in previous research (Rougeaux, Miranda, Fewtrell, & Wells, 2022). While the DHS provides nationally representative estimates of child growth outcomes between different maternal migration groups, they only include data for children under 5 years. At the time however, no data for older children were available.

Using the data from DHS Peru 2020, a mean height-for-age z score (HAZ) of -0.44 (SD: 0.79; *N*=2459) was found for children of urban non-migrant women and a mean HAZ of -0.77 (SD: 1.3; *N*=2586) for children of rural-urban migrant women. Calculations indicated a small effect size of 0.28. Using this data in G*Power indicated that, at an α of 0.05 and a power of 0.90 - both considered adequate values in health research for obtaining reliable findings - a sample of at least 428 would be needed to detect to detect a similar effect size in the Villa El Salvador study (Faul, 2007; Suresh & Chandrashekara, 2012). This is illustrated in the G*Power graph below (**Figure A**).

**Figure A.** G*Power output for comparison of child height growth means between two maternal internal migration groups using DHS for Peru 2020 (Faul 2007).

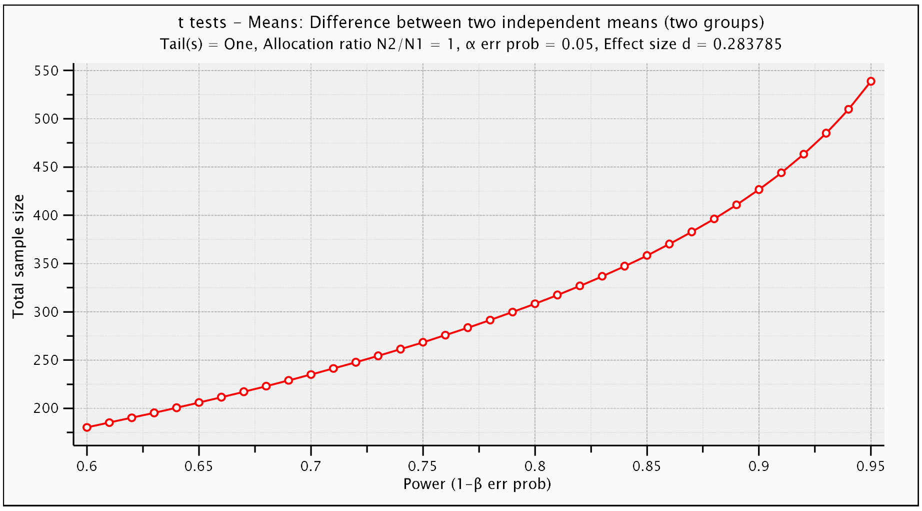

To allow sub-group analyses (such as assessing child outcomes by whether a child was born in a rural area versus an urban area), a decision was made to aim for a larger sample size of 600 of which half would be rural-urban migrant, a quarter urban non-migrant and a quarter urban-urban migrant mothers.

**Appendix S2: 12 item General Health Questionnaire (GHQ-12) questions and responses**

1. Have you recently, been able to concentrate on what you're doing? (Better than usual/Same as usual/Less than usual/Much less than usual)
2. Have you recently, lost much sleep over worry? (Not at all/More than usual/Rather more than usual/Much more than usual)
3. Have you recently, felt you were playing a useful part in things? (More so than usual/Same as usual/Less useful than usual/Much less than usual)
4. Have you recently, felt capable of making decisions about things? (More so than usual/Same as usual/Less so than usual/Much less capable)
5. Have you recently, felt constantly under strain? (Not at all/More than usual/Rather more than usual/Much more than usual)
6. Have you recently, felt you couldn't overcome your difficulties? (Not at all/More than usual/Rather more than usual/Much more than usual)
7. Have you recently, been able to enjoy your normal day-to-day activities? (More so than usual/Same as usual/Less so than usual/Much less than usual)
8. Have you recently, been able to face up to your problems? (More so than usual/Same as usual/Less so than usual/Much less able)
9. Have you recently, been feeling unhappy and depressed? (Not at all/No more than usual/Rather more than usual/Much more than usual)
10. Have you recently, been losing confidence in yourself? (Not at all/No more than usual/Rather more than usual/Much more than usual)
11. Have you recently, been thinking of yourself as a worthless person? (Not at all/No more than usual/Rather more than usual/Much more than usual)
12. Have you recently, been feeling reasonably happy, all things considered? (More so than usual/About same as usual/Less so than usual/Much less than usual)

Footnote: Respondents selected one response from those shown in parentheses for each question. The responses were scored in the order shown from 0 to 4, with a higher score indicating the poorest outcome. The scores were added up to give a total score ranging from 0 to 48, with a higher total score indicating greater distress (Goldberg et al., 1997).

### Appendix S3: Latent profile model approach for models with 1 to 6 classes

Models were compared using the Akaike Information Criterion (AIC), Bayesian Information Criterion (BIC), Lo–Mendell–Rubin (LMR) likelihood-ratio test and entropy statistics. Results of these comparisons are provided in the table below.

| Classes | AIC | BIC | LMR test statistic | LMR test p-value | Entropy |
|---|---|---|---|---|---|
| 1 | 16,618.93 | 16,660.56 | - | - | - |
| 2 | 16,255.13 | 16,321.74 | 365.90 | 0.039 | 0.7547 |
| 3 | 16,019.86 | 16,111.46 | 240.76 | 0.002 | 0.8469 |
| 4 | 15,868.15 | 15,984.72 | 159.4 | 0.574 | 0.8754 |
| 5 | 15,821.61 | 15,963.16 | 57.00 | 0.381 | 0.8436 |
| 6 | 15,774.25 | 15,940.79 | 57.80 | 0.409 | 0.8528 |

AIC = Akaike information criterion; BIC = Bayesian information criterion; LMR = Lo–Mendell–Rubin-adjusted likelihood-ratio.

Combinations of different options relating to the computation and selection of starting values, covariance of error terms, and variability of parameters across classes were also tested for each model, as recommended by Masyn (2013), and to take into account the underlying distributions of variables in the model.

A class invariant diagonal three-class model with robust standard errors was selected based on lower BIC and AIC values compared to other models and options, LMR p-values indicating appropriateness of a three-class model over others, a high entropy value closer to 1 (indicating higher classification accuracy), interpretability and non-normality of underlying data (Masyn 2013, Bauer 2022).

**Table S1:** Pearson pairwise correlations coefficients and p-values for biomarkers of maternal allostatic load

| | Body mass index | Waist circumference | Systolic blood pressure | Diastolic blood pressure | Cortisol (hair) |
|---|---|---|---|---|---|
| Body mass index | 1 | | | | |
| Waist circumference | 0.8618 | 1 | | | |
| | p=0.0000 | | | | |
| Systolic blood pressure | 0.3667 | 0.2873 | 1 | | |
| | p=0.0000 | p=0.0000 | | | |
| Diastolic blood pressure | 0.3066 | 0.2352 | 0.7563 | 1 | |
| | p=0.0000 | p=0.0000 | p=0.0000 | | |
| Cortisol (hair) | 0.0843 | 0.0904 | 0.1119 | 0.0526 | 1 |
| | p=0.0674 | p=0.0496 | p=0.015 | p=0.2543 | |

**Table S2:** Predicted means for the three-class latent profile model

| | Predicted mean | Standard error |
|---|---|---|
| Class 1: lower allostatic load | | |
| Body mass index | 25.4 | 0.3 |
| Waist circumference (cm) | 82.9 | 0.8 |
| Systolic blood pressure (mmHg) | 103.9 | 0.8 |
| Diastolic blood pressure (mmHg) | 61.7 | 0.6 |
| Cortisol (pg/mg) | 9.9 | 0.5 |
| Class 2: moderate allostatic load | | |
| Body mass index | 31.0 | 0.3 |
| Waist circumference (cm) | 96.8 | 0.7 |
| Systolic blood pressure (mmHg) | 111.2 | 0.8 |
| Diastolic blood pressure (mmHg) | 66.1 | 0.6 |
| Cortisol (pg/mg) | 10.5 | 0.4 |
| Class 3: higher allostatic load | | |
| Body mass index | 39.7 | 0.7 |
| Waist circumference (cm) | 113.0 | 1.4 |
| Systolic blood pressure (mmHg) | 118.7 | 2.5 |
| Diastolic blood pressure (mmHg) | 71.0 | 1.4 |
| Cortisol (pg/mg) | 11.0 | 1.3 |

**Table S3:** Pearson pairwise correlations coefficients and p-values for child markers of adiposity

| | BMI-for-age z score | Waist circumference-for-age z score |
|---|---|---|
| BMI-for-age z score | 1 | |
| Waist circumference-for-age z score | 0.8329 | 1 |
| | p=0.0000 | |

**Table S4:** Association of maternal mental distress and allostatic load, logistic regression odds ratios (95% CI; N=275)

| | Unadjusted | Adjusted for confounders† |
|---|---|---|
| Lower AL | Ref. | Ref. |
| Moderate AL | 2.07 (1.26, 3.40)* | 2.02 (1.21, 3.38)* |
| Higher AL | 1.48 (0.64, 3.42) | 1.42 (0.60, 3.37) |

† Confounders = child sex, age, maternal age, migration status, ethnicity, and years lived in an urban area; * p<0.05; OR=odds ratio, CI=confidence interval; maternal mental distress score was obtained from the 12-item General Health Questionnaire and dichotomised as scores of 5 and over indicating moderate-high mental distress versus no-low mental distress (reference).

**Table S5** Associations of individual items of the 12-item General Health Questionnaire and child adiposity markers, linear regression coefficients (95% CI; N=275)

| | Unadjusted (univariate) | Adjusted for all items & confounders† |
|---|---|---|
| **BMI-for-age z score (WHO)** | | |
| 1. Have you recently, been able to concentrate on what you're doing? | -0.14 (-0.38, 0.10) | -0.11 (-0.35, 0.13) |
| 2. Have you recently, lost much sleep over worry? | -0.02 (-0.26, 0.21) | -0.05 (-0.28, 0.19) |
| 3. Have you recently, felt you were playing a useful part in things? | -0.19 (-0.61, 0.24) | -0.15 (-0.57, 0.27) |
| 4. Have you recently, felt capable of making decisions about things? | -0.29 (-0.66, 0.08) | -0.25 (-0.61, 0.12) |
| 5. Have you recently, felt constantly under strain? | -0.11 (-0.34, 0.13) | -0.11 (-0.34, 0.13) |
| 6. Have you recently, felt you couldn't overcome your difficulties? | -0.29 (-0.57, -0.02)* | -0.23 (-0.50, 0.05) |
| 7. Have you recently, been able to enjoy your normal day-to-day activities? | -0.02 (-0.34, 0.29) | 0.00 (-0.32, 0.31) |
| 8. Have you recently, been able to face up to your problems? | -0.49 (-0.90, -0.07)* | -0.42 (-0.84, -0.01)* |
| 9. Have you recently, been feeling unhappy and depressed? | -0.12 (-0.38, 0.13) | -0.11 (-0.36, 0.14) |
| 10. Have you recently, been losing confidence in yourself? | -0.36 (-0.70, -0.01)* | -0.32 (-0.66, 0.03) |
| 11. Have you recently, been thinking of yourself as a worthless person? | -0.85 (-1.30, -0.39)* | -0.68 (-1.15, -0.22)* |
| 12. Have you recently, been feeling reasonably happy, all things considered? | -0.37 (-0.73, 0.00) | -0.33 (-0.69, 0.03) |
| **Waist circumference-for-age z score (internal)** | | |
| 1. Have you recently, been able to concentrate on what you're doing? | -0.05 (-0.24, 0.13) | -0.07 (-0.25, 0.12) |
| 2. Have you recently, lost much sleep over worry? | -0.04 (-0.22, 0.14) | -0.07 (-0.25, 0.12) |
| 3. Have you recently, felt you were playing a useful part in things? | -0.08 (-0.40, 0.25) | -0.05 (-0.37, 0.28) |
| 4. Have you recently, felt capable of making decisions about things? | -0.18 (-0.46, 0.11) | -0.18 (-0.46, 0.11) |
| 5. Have you recently, felt constantly under strain? | -0.06 (-0.24, 0.12) | -0.09 (-0.27, 0.10) |
| 6. Have you recently, felt you couldn't overcome your difficulties? | -0.15 (-0.36, 0.07) | -0.12 (-0.33, 0.10) |
| 7. Have you recently, been able to enjoy your normal day-to-day activities? | -0.04 (-0.28, 0.21) | -0.04 (-0.28, 0.20) |
| 8. Have you recently, been able to face up to your problems? | -0.28 (-0.60, 0.04) | -0.28 (-0.60, 0.04) |
| 9. Have you recently, been feeling unhappy and depressed? | -0.11 (-0.31, 0.08) | -0.12 (-0.31, 0.08) |
| 10. Have you recently, been losing confidence in yourself? | -0.27 (-0.54, -0.01)* | -0.25 (-0.52, 0.02) |
| 11. Have you recently, been thinking of yourself as a worthless person? | -0.64 (-1.00, -0.29)* | -0.60 (-0.93, -0.21)* |
| 12. Have you recently, been feeling reasonably happy, all things considered? | -0.21 (-0.49, 0.07) | -0.20 (-0.49, 0.08) |

† Confounders = child sex, age, maternal age, migration status, ethnicity, and years lived in an urban area; * p<0.05; BMI=body mass index, WHO=World Health Organization, CI=confidence interval.

**Table S6** Associations of individual biomarkers of allostatic load and child adiposity markers, linear regression coefficients (95% CI; N=275)

| | Unadjusted univariate | Adjusted for all allostatic load biomarkers | Adjusted for allostatic load biomarkers & confounders† |
|---|---|---|---|
| **Child BMI-for-age z-score (WHO)** | | | |
| Maternal cortisol (log pg/mg) | 0.16 (-0.06, 0.38) | 0.12 (-0.09, 0.33) | 0.18 (-0.02, 0.39) |
| Maternal BMI (kg.m$^2$) | 0.09 (0.07, 0.11)* | 0.08 (0.04, 0.13)* | 0.07 (0.03, 0.12)* |
| Maternal waist circumference (cm) | 0.03 (0.02, 0.04)* | 0.00 (-0.02, 0.02) | 0.01 (-0.01, 0.03) |
| Maternal systolic blood pressure (mmHg) | 0.01 (0.00, 0.02)* | -0.01 (-0.02, 0.01) | 0.00 (-0.01, 0.02) |
| Maternal diastolic blood pressure (mmHg) | 0.02 (0.01, 0.03)* | 0.01 (-0.01, 0.03) | 0.01 (-0.02, 0.03) |
| **Child waist circumference-for-age z score (internal)** | | | |
| Maternal cortisol (log pg/mg) | 0.15 (-0.02, 0.32) | 0.11 (-0.05, 0.27) | 0.14 (-0.02, 0.30) |
| Maternal BMI (kg.m$^2$) | 0.06 (0.04, 0.08)* | 0.00 (-0.03, 0.04) | 0.00 (-0.04, 0.03) |
| Maternal waist circumference (cm) | 0.03 (0.02, 0.04)* | 0.03 (0.01, 0.04)* | 0.03 (0.01, 0.05)* |
| Maternal systolic blood pressure (mmHg) | 0.01 (0.01, 0.02)* | 0.00 (-0.01, 0.02) | 0.01 (-0.01, 0.02) |
| Maternal diastolic blood pressure (mmHg) | 0.02 (0.00, 0.03)* | 0.00 (-0.01, 0.02) | 0.00 (-0.02, 0.02) |

† Confounders = child sex, age, maternal age, migration status, ethnicity, and years lived in an urban area; * $p<0.05$; BMI=body mass index, WHO=World Health Organization, CI=confidence interval.

**Figure S1.** Conceptual diagram showing hypothesized associations of two maternal exposures of stress (as mental distress and allostatic load) and two child adiposity markers (BMI and waist circumference).

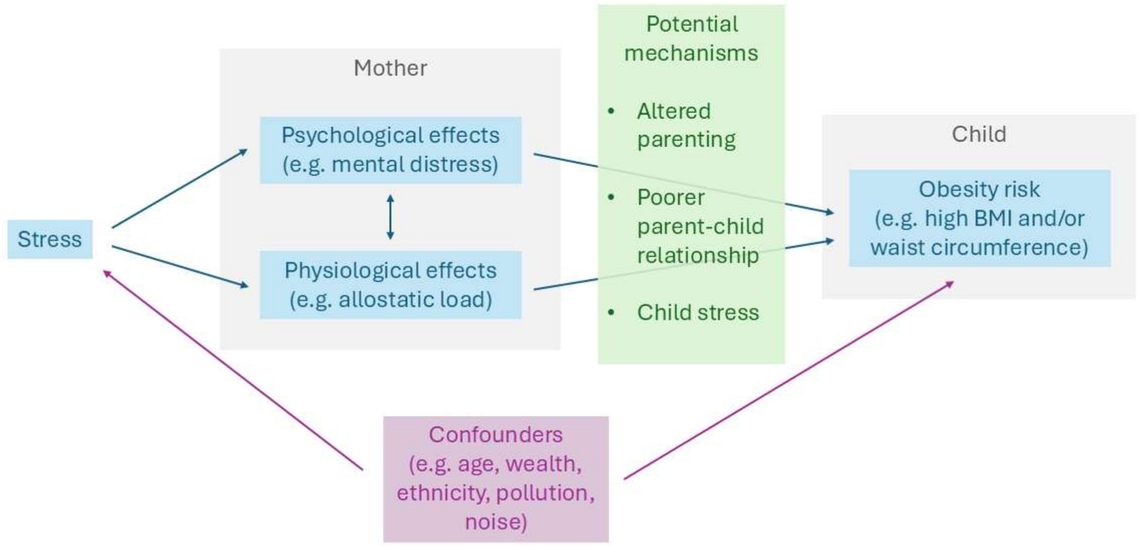
